# International Collaborative Study on Human Papillomavirus Analytical Thresholds for Sensitivity and Specificity in Cervical Screening

**DOI:** 10.64898/2026.02.03.26345438

**Authors:** Emel Yilmaz, Gerald L. Murray, Prisha Balgovind, Suzanne M. Garland, A. Rita Pereira, Davy Vanden Broeck, Nina Redzic, Jean-Luc Prétet, Quentin Lepiller, Steffi Silling, Clementina Cocuzza, Marianna Martinelli, Allan Campbell, Conor Brown, Kate Cuschieri, Linzi Connor, Anja Oštrbenk, Mario Poljak, Murat Gultekin, Yalın Kılıç, K. Miriam Elfström, Laila Sara Arroyo Mühr, Joakim Dillner

## Abstract

**Background:** Cervical screening using human papillomavirus (HPV) testing is a pillar of global cervical cancer elimination. However, different HPV assays vary in both the HPV types they detect as well as the minimum amount of virus they detect. The aim of this collaborative study was to define which HPV type-specific analytical threshold of detection provides optimal sensitivity and specificity of cervical screening.

**Methods:** 100 cervical intraepithelial neoplasia grade 2 or worse (CIN2+) cases and 200 matched population-based controls were obtained at the Swedish National HPV Reference Laboratory and analyzed by 10 laboratories across 10 countries. Cumulative sensitivity (weighted according to the global HPV type distribution in invasive cervical cancer (ICC)) and specificity were estimated at varying analytical detection thresholds.

**Results:** Consensus results found HPV in 99/100 CIN2+ cases and 52/200 controls. HPV16 prevalence declined in HPV-vaccinated birth cohorts, among both cases and controls. Line plots of 1-specificity and ICC-weighted sensitivity found optimal analytical detection thresholds as 3 International Units (IU)/µl for HPV16/18, 25 IU/µl for HPV31/33/35/45/52/58 and 100 genome equivalents/µl of HPV 39/51/56/59 resulting in 92.00% cumulative specificity and 90.08% ICC-weighted sensitivity.

**Conclusion:** An international collaborative study has identified HPV analytical detection thresholds optimizing sensitivity and specificity of cervical screening.

## INTRODUCTION

Cervical screening based on human papillomavirus (HPV) testing is more sensitive and objective than cytology in preventing invasive cervical cancer (ICC) (1). The World Health Organization (WHO) has named high-performance HPV-based screening as a pillar of the global strategy to eliminate cervical cancer (2).

High amounts of HPV DNA are associated with infection persistence, with low amounts linked to transient infections (3, 4). High HPV amounts may also be associated with progression to cervical intraepithelial neoplasia grade 2 or worse (CIN2+) (5–9). This has been consistently observed for HPV16 (10–12). A prospective cohort study with ICC as endpoint, reported optimal sensitivity and specificity when HPV16/18 were detected at low quantities, but higher analytical thresholds were used for HPV 31/33/45/52 (13). Although early ICC detection is one purpose of screening, the main purpose is identifying and treating CIN2+ lesions.

As of December 2023, more than 260 different HPV tests are commercially available (in addition to numerous in-house methods) (14). Performances vary, both by assay and by the laboratory performing the tests (15–24). Criteria for validation of HPV assays have focused on clinical sensitivity and specificity for detection of CIN2+, without consideration of the amount of virus that should be detectable. We launched an international collaborative study involving expert laboratories from multiple countries, including national HPV reference laboratories (NRL). An international consensus on required analytical thresholds for HPV assays would be important for quality assurance and continued development and validation of HPV assays.

## MATERIAL AND METHODS

The Swedish cervical screening program mandates HPV-based screening for all women aged 23-70 years and used reflex cytology after HPV-positivity. Women with cytological abnormalities are referred for cervical biopsy and histopathology. Cases and controls were identified by the Center for Cervical Cancer Elimination (CCCE), the central laboratory for the regional population-based screening program. CCCE registers the HPV tests, cytology, and histopathology results. Cases were women with CIN2+ in histopathology with a liquid-based cytology (LBC) sample registered at or at most three months before CIN2+ diagnosis. Controls were women participating in the population-based screening program who had LBC samples taken for primary screening but no CIN2+ in histopathology during the same follow-up period as the cases. For each case, two controls were selected, matched by age (±5 years). 100 consecutive eligible cases and 200 age matched controls were identified between 4^th^ April 2022 and 27^th^ March 2023. The mean ages were 34.33 years (cases) and 35.40 years (controls).

We prepared dilution series of HPV DNA (whole genome HPV plasmids) for each of the 12 HPV types that are classified as oncogenic by the International Agency for Research on Cancer (IARC): HPV16/18/31/33/35/39/45/51/52/56/58/59 (25). For HPV16/18/31/33/45/52/58 we used International Standards (IS) of HPV DNA and report concentrations in International Units (IU)/µl. For HPV35/39/51/56/59, dilution series used defined amounts of genome equivalents (GE)/µl of HPV DNA. The ten-fold dilutions ranged from 10^5^ to 10^0^ IU or GE per µl, yielding a total of 72 standards for calibration. HPV DNA was diluted in TE buffer (10 mM TRIS-HCL, 0.1 mM EDTA, pH 8.0) containing 10 ng/µl human DNA. Study sample aliquots and standards were distributed to expert laboratories in 10 countries participating in the global HPV LabNet (Australia, Belgium, France, Germany, Italy, Scotland, Slovenia, Sweden, Türkiye and USA). Each aliquot was anonymized with unique identifier and shipped blinded, without any clinical, pathological or virological information.

The participating laboratories performed analyses using their extraction and HPV testing methods. NRL Sweden used Cobas 4800 (Roche Diagnostics, Basel, Switzerland) or BD Onclarity™ HPV Assay (Becton, Dickinson and Company, Eysins, Switzerland) for the cervical screening program. The reference method for HPV quantification used the Hamilton® Microlab Star (Hamilton Bonaduz AG, Bonaduz, Switzerland) automated DNA extraction method with the Mag-Bind® Universal Pathogen 4×96 (Omega Bio-Tek, Leverkusen, Germany) kit and HPV analysis on the Quant Studio™ 3 Real-Time PCR system (Thermo Fisher Scientific Applied Biosystems™, Darmstadt, Germany), targeting the E6/E7 region of HPV16, 18, 31, 33, 35, 39, 45, 51, 52, 56, 58, 59, respectively Real-time PCR reactions in 20 µL volumes contained 50-100 ng of genomic DNA, GoTaq® Probe Master Mix (Promega Corporation, Mannheim, Germany), 0.5 µM of each forward and reverse primer, and 0.25 µM of HPV QSY-probe. Standards were run as triplicates while study samples were run as duplicates and the mean amounts of HPV recorded.

The NRL of Australia used Roche MagNA Pure 96 DNA (Roche Diagnostics, Basel, Switzerland) with Viral NA Small Volume Kit (Roche Diagnostics, Basel, Switzerland) for DNA extraction and performed HPV detection using Anyplex™ II HPV HR Detection (Seegene Inc, Seoul, South Korea) and quantitation on an in-house droplet digital PCR targeting the E6 gene on the Bio-Rad QX100/QX200 system (Bio-Rad Laboratories, California, USA) (manuscript in preparation); NRL of Belgium used Chemagic 360 system (Revvity, Turku, Finland) for automated DNA extraction from liquid-based cytology samples, employing magnetic bead-based technology and for HPV analysis the in-house developed RIATOL quantitative real-time PCR genotyping assay (AML Laboratory, Antwerp, Belgium) that individually targets 18 HPV genotypes (HPV6, 11, 16, 18, 31, 33, 35, 39, 45, 51, 52, 53, 56, 58, 59, 66, 67, and 68) using eight multiplex reactions per sample was performed at ultra-low reaction volumes on the LightCycler instrument (Roche Diagnostics, Basel, Switzerland); NRL of Germany used Roche Magna Pure DNA/Viral NA LV 2.0 (Roche Diagnostics, Basel, Switzerland) for extracting DNA from the samples and performed HPV analysis using Allplex™ HPV28 Detection platform (Seegene Inc, Seoul, South Korea) as well as HPV type-specific in-house real time PCR; NRL of France used Microlab Nimbus (Hamilton Bonaduz AG, Bonaduz, Switzerland) with the NucleoMag Dx Pathogen kit (Macherey Nagel, Düren, Germany) for DNA extraction and in-house real-time PCR assay for HPV analysis; NRL Italy used Microlab Nimbus for DNA extraction and OncoPredict HPV QT assay (Hiantis, Milan, Italy) for HPV analysis; NRL of Scotland performed DNA extraction using Universal extraction system with STARMag Universal Cartridge kit (Seegene Inc, Seoul, South Korea) and conducted HPV analysis completed HPV analysis using Allplex™ HPV28 Detection platform; NRL of Slovenia used QIAamp DNA Mini Kit (Qiagen, Hilden, Germany) for DNA extraction and Allplex™ HPV HR for HPV testing; NRL USA analyzed the samples for HPV using Typeseq2 (Frederick National Laboratory for Cancer Research, Maryland, United States);participating laboratory from Türkiye used in-house DNA isolation kit (DNA Elixir Smear, Izmir, Türkiye) to extract DNA from the samples and in-house real-time PCR assay (CarciScan HPV PCR Assay, Izmir, Türkiye) for HPV analysis.

The 10 countries provided 13 datasets, as some laboratories used >1 assay. Germany and Sweden performed virus quantification on HPV-positive samples. For consensus on presence of HPV, we required ≥70% agreement regarding the detected HPV type. If multiple HPV types were detected in the same sample, positivity was assigned following hierarchy of cancer risk among class I carcinogen viruses (25): HPV16>18>45>33>58>31>52>35>59>39>51>56>other HPV types. Several labs also reported viruses not established as oncogenic: HPV6/40/42/44/54/61/66/68/70/82 which are here termed as “Other HPV”. Cumulative sensitivity and specificity for detecting a case were first calculated for each HPV type with all reported interpretation as positive even if they corresponded to extremely low virus amounts.

Cycle threshold (Ct) values from serially diluted standards, were utilized to generate standard curves for HPV types 16/18/31/33/35/39/45/51/52/56/58/59. Two laboratories did not detect standards at the lowest concentrations and only data from five ten-fold dilutions (10^5^ – 10^1^ IU or GE per µl) were included from their datasets. Some laboratories reported additional HPV types that should not be present in the standards. A previously reported contamination with low amounts of HPV35 in the HPV31 standard was confirmed (24), whereas all other false positives were limited to the reporting laboratory.

Seven laboratories provided Ct values for both calibration and clinical samples, enabling quantification of viral amounts using standard curves. Theoretical standard concentrations were log-transformed, and linear regression performed with Ct values as the dependent variable and log10 concentrations as the independent variable, yielding slope, intercept, and R-squared values. White’s test was used to assess heteroscedasticity. HPV type-specific virus amounts in each HPV-positive sample were calculated as 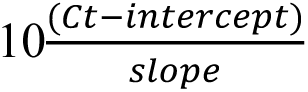 (26, 27). Virus amounts per µl of original sample volume were calculated based on the input sample volume in extraction and volume of the resulting extracted DNA material. For each HPV type, median viral amounts were determined for datasets that were positive for the consensus virus. As many CIN lesions are caused by HPV types with limited contribution to ICC, sensitivities were weighted according to the importance of each virus in the etiology of ICC using global attributable fractions (25). Cumulative sensitivity, cumulative specificity, and ICC-weighted sensitivity were calculated. Line plots evaluated relationship between 1-specificity and ICC-weighted sensitivity across varying analytical detection thresholds, by HPV amounts/µl of original sample volume (Figure 1). All statistical analyses used STATA, version 17. R version 4.3.2 generated the figure and for data transpose.

**Figure 1:**
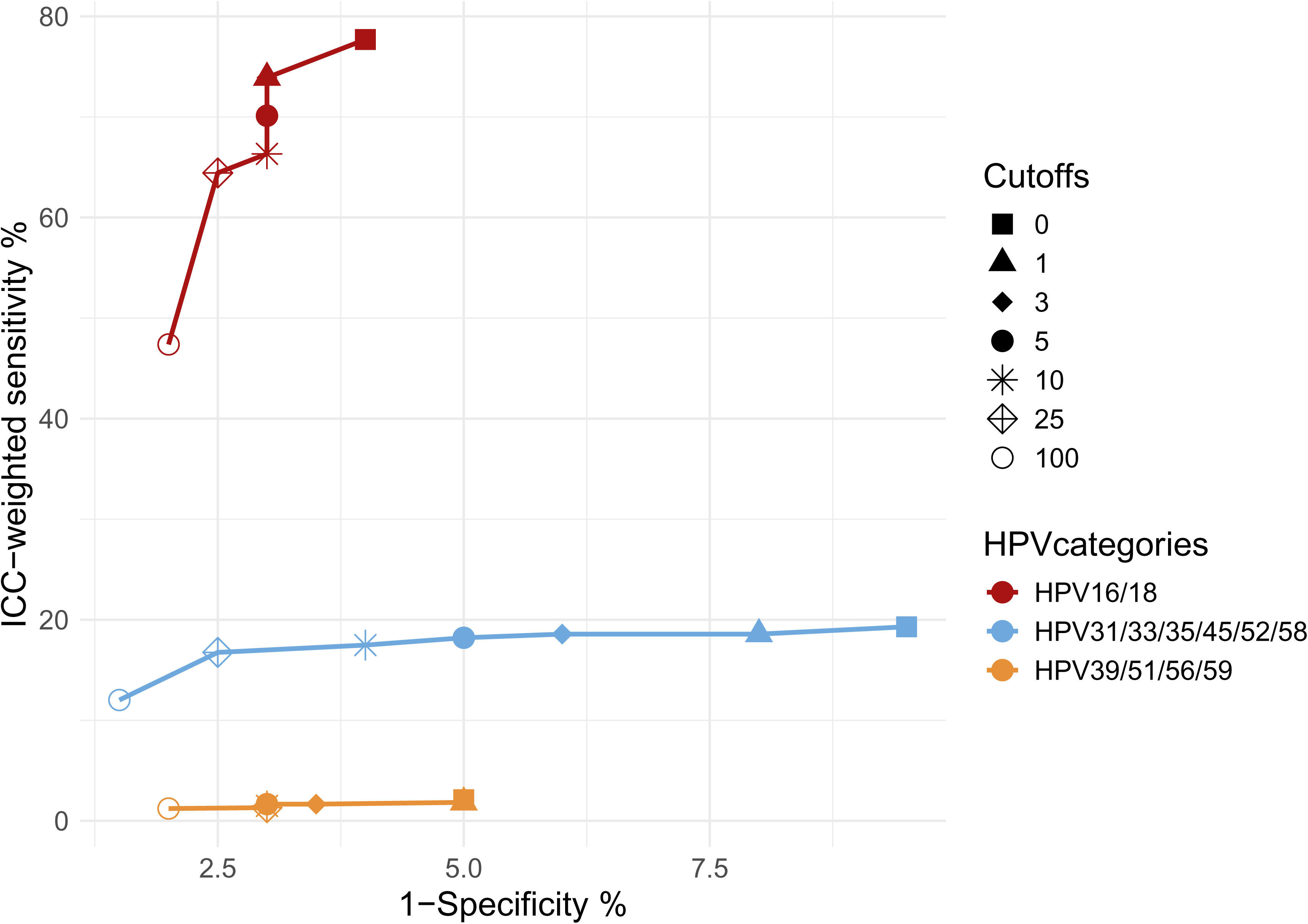
Line plot comparing ICC weighted sensitivity and 1-specificity based on varying thresholds per µl of sample volume for detection of three main categories of HPV types.

Ethical approval was granted by the national ethical review agency of Sweden (DNR2023-02457-01, DNR2025-08540-02). The ethical review agency determined that informed consent was not required.

## RESULTS

In the routine primary HPV screening (using BD Onclarity and Cobas 4800), 99/100 cases (99.00%) and 37/200 controls (18.50%) were reported as HPV-positive. The Cobas 4800 assay reported results only in three channels (HPV16, HPV18 and non-16/18 oncogenic HPV). Samples reported as positive for non-16/18 oncogenic HPV were re-analysed using BD Onclarity for extended genotyping, of those two samples were HPV negative in BD. Reflex cytologies were abnormal for 94/100 cases. Among controls, there were eight abnormal and 31 normal cytologies. For 161 controls, there was no cytology (because of HPV negativity). The histopathologies among cases were 94 high-grade squamous intraepithelial lesions (HSIL), two adenocarcinomas *in situ* (AIS), two AIS/HSIL and two squamous cell carcinomas (SCC). Among controls, there were two low-grade squamous intraepithelial lesions (LSIL) and six normal histopathologies (remaining controls had no biopsy).

Consensus results from 13 datasets found HPV in 52/200 (26.00%) controls and 99/100 (99.00%) cases (all signals were considered including detection of possibly/probably or not oncogenic HPV types) resulting in cumulative sensitivity and specificity of 99.00% and 74.00%, respectively (Table 1). Stratifying by birth cohorts with or without organized catch-up vaccination with quadrivalent HPV vaccine (HPV6/11/16/18; population coverage ∼55%) (28), vaccinated birth cohorts had 6/32 HPV16 positive cases (18.75%), compared with 28/68 cases (41.18%) among women born 1954-1962 (p=0.041, Fischer’s exact test among cases).

**Table 1:** Consensus on reference HPV type distribution by cases and controls determined from 13 datasets from 10 participating laboratories. Thresholds for virus copies were not applied; the interpretation of the testing assays was approved in case of raw data all signals were accepted as positive for HPV. In case of positivity for multiple HPV types, the most oncogenic HPV type is listed.

| HPV type | Total | Control | Case | Cumulative sensitivity% | Cumulative Specificity (%) |
| --- | --- | --- | --- | --- | --- |
| 16 | 40 | 6 | 34 | 34.00 | 97.00 |
| 18 | 9 | 2 | 7 | 41.00 | 96.00 |
| 45 | 6 | 2 | 4 | 45.00 | 95.00 |
| 33 | 12 | 3 | 9 | 54.00 | 93.50 |
| 58 | 13 | 7 | 6 | 60.00 | 90.00 |
| 31 | 20 | 5 | 15 | 75.00 | 87.50 |
| 52 | 10 | 2 | 8 | 83.00 | 86.50 |
| 35 | 6 | 1 | 5 | 88.00 | 86.00 |
| 59 | 3 | 1 | 2 | 90.00 | 85.50 |
| 39 | 6 | 3 | 3 | 93.00 | 84.00 |
| 51 | 3 | 2 | 1 | 94.00 | 83.00 |
| 56 | 6 | 3 | 3 | 97.00 | 81.50 |
| Other HPV* | 17 | 15 | 2 | 99.00 | 74.00 |
| Not detectable/No consensus | 149 | 148 | 1 | 100.00 | 0.00 |
| Total | 300 | 200 | 100 |  |  |

\*HPV 6, 40, 42, 44, (44 & 61), 54, 61, 66, 68, (68, 70), 82.

Line plot inspection (Figure 1) found that 3 IU/µl for HPV16 and 18 was required for adequate sensitivity and that 25 IU/µl for HPV31/33/35/45/52/58 and 25 GE/µl for HPV35 gave an adequate sensitivity with limited adverse effect on specificity. For the lower oncogenicity HPV types (HPV39/51/56/59) their joint contribution to sensitivity was minimal, whereas detection at low thresholds gave a strong adverse effect on specificity (Figure 1). Using a threshold of 100 GE/ul for HPV39/51/56/59 improved specificity.

Table 2 compares the consensus HPV distribution in cases and controls with the HPV-positives remaining after applying analytical thresholds. The number of HPV-positives was reduced in particular among controls, increasing specificity. ICC-weighted sensitivity of the major oncogenic HPV types 16/18/31/33/35/45/52/58 jointly was 88.72% (Table 2). When detection of HPV39/51/56/59 was included, ICC-weighted sensitivity was 90.08% with a cumulative specificity of 92.00% (Table 2).

**Table 2:** Consensus HPV type distribution among 7 participating laboratories by cases and controls. with applied thresholds at 3 IU/ µl of sample volume for HPV16 and 18; 25 IU/µl of sample volume for HPV31, 33, 35, 45, 52 and 58; 25 GE/ ul of sample volume for HPV35, 100 GE/ µl of sample volume for HPV39, 51, 56 and 59. In case of positivity for multiple HPV types, the most oncogenic HPV type is listed. ICC weighted sensitivity was calculated using population attributable fraction (PAF) adapted from IARC (25).

| HPV type | Controls -<br>Reference | Controls –<br>with<br>thresholds | Cases -<br>reference | Cases -<br>with<br>thresholds | Controls<br>remained<br>positive<br>after<br>thresholds | Cases<br>remained<br>positive after<br>thresholds | Sensitivity** | Cumulative<br>sensitivity%*** | Cumulative<br>Specificity%<br>*** | PAF<br>ICC% | Sensitivity<br>with<br>threshold% | Cumulative<br>Sensitivity<br>ICC<br>weighted% |
| --- | --- | --- | --- | --- | --- | --- | --- | --- | --- | --- | --- | --- |
| 16 | 6 | 4 | 34 | 33 | 4 | 33 | 0.97 | 33.00 | 98.00 | 62.40 | 60.56 | 60.56 |
| 18 | 2 | 2 | 7 | 6 | 2 | 6 | 0.86 | 39.00 | 97.00 | 15.30 | 13.11 | 73.68 |
| 45 | 2 | 1 | 4 | 3 | 1 | 3 | 0.75 | 42.00 | 96.50 | 4.80 | 3.60 | 77.28 |
| 33 | 3 | 0 | 9 | 8 | 0 | 8 | 0.89 | 50.00 | 96.50 | 3.90 | 3.47 | 80.75 |
| 58 | 7 | 1 | 6 | 5 | 1 | 5 | 0.83 | 55.00 | 96.00 | 3.70 | 3.08 | 83.83 |
| 31 | 5 | 3 | 15 | 14 | 3 | 13 | 0.87 | 69.00 | 94.50 | 2.90 | 2.51 | 86.34 |
| 52 | 2 | 0 | 8 | 5 | 0 | 3 | 0.38 | 74.00 | 94.50 | 2.60 | 0.98 | 87.32 |
| 35 | 1 | 0 | 5 | 7 | 0 | 5 | 1.00 | 81.00 | 94.50 | 1.40 | 1.40 | 88.72 |
| 59 | 1 | 1 | 2 | 2 | 1 | 2 | 1.00 | 83.00 | 94.00 | 0.90 | 0.90 | 89.62 |
| 39 | 3 | 1 | 3 | 1 | 1 | 1 | 0.33 | 84.00 | 93.50 | 0.80 | 0.27 | 89.88 |
| 51 | 2 | 1 | 1 | 2 | 0 | 0 | 0.00 | 86.00 | 93.00 | 0.20 | 0.00 | 89.88 |
| 56 | 3 | 2 | 3 | 3 | 1 | 3 | 1.00 | 89.00 | 92.00 | 0.20 | 0.20 | 90.08 |
| Other HPV* | 15 | - | 2 | - | - | - | - | 89.00 | 92.00 | - | - | - |
| Neg/Not<br>detectable/No<br>consensus | 148 | 184 | 1 | 11 | 148 | 1 |  | 100.00 | 0.00 |  |  |  |

\*HPV 6, 40, 42, 44, (44 & 61), 54, 61, 66, 68, (68, 70), 82.
\*\*Calculated as: Cases remained positive after thresholds / Cases - reference
\*\*\* Based on HPV distribution among cases and controls after thresholds were applied.

A few very well characterized HPV assays are considered as comparator assays in validation studies, including Cobas 4800 and BD Onclarity (referred to as BD/Cobas) (29). Table 3 compares the consensus HPV distribution in cases and controls with the HPV-positives for the same types from BD/Cobas. The comparator assay testing using manufacturer-defined thresholds achieved high ICC-weighted sensitivity (96.61%) but much lower specificity (82.50%).

**Table 3:** HPV type distribution determined by BD/Cobas with consensus HPV distribution from Table 1 as reference. In case of positivity for multiple HPV types, the most oncogenic HPV type is listed. ICC weighted sensitivity was calculated using PAF adapted from IARC (25).

| HPV type | Control -<br>consensus | Controls<br>–<br>BD/Cobas | Case -<br>consensus | Case-BD/Cobas | Controls -<br>BD/Cobas<br>positives<br>when<br>consensus<br>positive | Cases<br>BD/Cobas<br>positives<br>when<br>consensus<br>positive | Sensitivity** | Cumulative<br>sensitivity%*** | Cumulative<br>Specificity%*** | PAF<br>ICC% | Sensitivity<br>with<br>threshold% | Cumulative<br>Sensitivity<br>ICC<br>weighted% |
| --- | --- | --- | --- | --- | --- | --- | --- | --- | --- | --- | --- | --- |
| 16 | 6 | 10 | 34 | 34 | 6 | 34 | 1.00 | 34.00 | 95.00 | 62.40 | 62.40 | 62.40 |
| 18 | 2 | 2 | 7 | 6 | 2 | 6 | 0.86 | 40.00 | 94.00 | 15.30 | 13.11 | 75.51 |
| 45 | 2 | 1 | 4 | 4 | 1 | 4 | 1.00 | 44.00 | 93.50 | 4.80 | 4.80 | 80.31 |
| P1 (33/58) | 10 | 8 | 15 | 14 | 8 | 14 | 0.93 | 58.00 | 89.50 | 7.60 | 7.09 | 87.41 |
| 31 | 5 | 4 | 15 | 15 | 4 | 15 | 1.00 | 73.00 | 87.50 | 2.90 | 2.90 | 90.31 |
| 52 | 2 | 1 | 8 | 8 | 1 | 8 | 1.00 | 81.00 | 87.00 | 2.60 | 2.60 | 92.91 |
| P3 (35/39/68) | 7 | 4 | 9 | 11 | 4 | 9 | 1.00 | 92.00 | 85.00 | 2.40 | 2.40 | 95.31 |
| P2 (56/59/66) | 6 | 5 | 6 | 6 | 5 | 6 | 1.00 | 98.00 | 82.50 | 1.10 | 1.10 | 96.41 |
| 51 | 1 | 0 | 1 | 1 | 0 | 1 | 1.00 | 99.00 | 82.50 | 0.20 | 0.20 | 96.61 |
| Other HPV* | 11 | - | 0 | - | - | - | - | 99.00 | 82.50 | - | - | - |
| Neg/Not<br>detectable/No<br>consensus | 148 | 165 | 1 | 1 | 148 | 1 | 1.00 | 100.00 | 0.00 |  |  |  |

\*HPV 6, 40, 42, 44, 61, (44 & 61), 54, 61, 66, 68, (68, 70), 82. \*\*Calculated as: Cases remained positive after thresholds / Cases - reference \*\*\*Calculated based BD/Cobas output on HPV distribution among cases.

## DISCUSSION

We explored optimal analytical detection thresholds of individual HPV types in HPV tests intended for cervical screening. Because even reference laboratory testing can show considerable variability, an international collaborative study with 10 participating countries was conducted using the same samples. The identified type-specific analytical detection thresholds for the 12 oncogenic HPV types 16/18/31/33/35/39/45/51/52/56/58/59 could be used to greatly improve specificity (from 81.50% to 92.00%) with marginal loss of sensitivity. When analysis was restricted to the 8 major oncogenic HPV types (16/18/31/33/35/45/52/58), specificity increased further to 94.50%. Given that HPV-based screening is now a globally recommended public health policy, these improvements in specificity would correspond to >10% of the global screening-eligible population of women no longer being classified as screen-positive, while potentially missing only a small proportion of the CIN2+ lesions with potential to progress to ICC.

A strength of this study is its international robustness, combining results from 10 countries/laboratories using several different assay platforms. Additionally, study samples were selected from a population-based screening program, ensuring robust contextual data while both semi-quantitative and quantitative outputs enabled comprehensive analysis. Furthermore, expressing results in IU, traceable to WHO IS, ensures global comparability, both with current and future testing systems. However, several limitations should be noted. The study sample size was limited, resulting in a small number of observations for uncommon HPV types. Also, different screening programs may use different sample-taking systems. The LBC samples used in the study (collected with cervical brush suspended in 20 ml ThinPrep medium) represent the most common sampling method today, but alternative sampling media and collection systems exist. Analytical thresholds should be able to accommodate also future changes in how the samples are collected. Most screening tests do not relate the HPV test results to the amounts of cells in the sample, but in order for the thresholds to be transferable across sampling systems, we also provide data on how they perform if the amount of HPV is expressed in relation to the number of human cells in the sample.

The WHO target product profile (TPP) outlines principles for HPV tests for screening that are appropriate for public health needs (30). The TPP specifies high-performance HPV tests that detect, as a minimum, types 16/18/31/33/35/45/52/58 and specifically recommend against including HPV types outside of the 12 HPV types formally established as oncogenic (such as HPV68 and 66). For absolute clinical sensitivity of CIN2+, a minimum standard of 90% was proposed. TPP also highlighted that, although current HPV tests generally achieve very high sensitivity, specificity remains suboptimal. Our findings align with the TPP: both reference laboratory testing without thresholds and comparator assay testing had very high sensitivity, but suboptimal specificity. Including HPV types not formally classified as oncogenic greatly reduced specificity (to 74.00%), with negligible gains in sensitivity.

Older validation methods are dependent on clinical samples with longitudinal follow up for disease, making validation slow and expensive. Furthermore, the most oncogenic HPV types are becoming less common in CIN2+ in vaccinated birth cohorts. Present-day series of CIN2+ will contain non-vaccine HPV types of limited relevance for cervical cancer. Indeed, we found a decline of HPV16 in CIN2+ among birth cohorts that had been offered vaccination, necessitating weighting of the sensitivity according to the importance of the different HPV types for invasive cervical cancer, as assessed in the era before widespread HPV vaccination.

Many HPV tests define a threshold for HPV positivity using Ct values from a PCR reaction. However, different PCR-based tests amplify at different efficiencies and Ct values from one assay cannot be translated to Ct values for another. Also, Ct values are not linearly related to the amount of virus. Calculating the amount of virus using a standard curve of samples with defined amounts of virus in IU reduces these problems and increases international comparability. Since nearly all assays used in this study detected HPV types separately, threshold values for detection could be applied to a single HPV type.

This study included only clinician-taken samples. Assessment of optimal thresholds for detection in self-collected samples would require an entirely new study as the converted Ct outputs for different virus concentrations may differ from those of samples taken by healthcare personnel. Furthermore, pre-analytical variations in laboratory workflow such as the extraction method, the suspension volume of the analysis platforms, and operator issues with sample collection may also affect the virus amount in samples. To minimize accuracy and precision errors, standards should be analyzed multiple times to determine median Ct values and estimate virus quantities in samples in relation to standard curves.

The current study is a first step towards defining virological endpoints for evaluation of HPV screening tests. The limited sample size was a necessity given the large number of participating laboratories. In further exploration of virological endpoints, many different settings are likely to be able to investigate large laboratory databases from real-life screening programs which would provide data on the robustness of the required analytical threshold of detection for HPV type specific virus amounts proposed here.

In conclusion, this study defined analytical detection thresholds for HPV genotypes that achieved much higher specificity while maintaining adequate sensitivity in HPV-based cervical screening. Definition of which amount of virus, traceable to IS, should be detectable for optimal sensitivity and specificity in cervical screening could facilitate the development, validation and quality assurance of HPV testing methods.

## ACKNOWLEDGEMENTS

We would like to thank Alice Baraquin and Killian Jacquot (French HPV NRL) for excellent technical assistance with HPV quantification and Elisabeth R. Unger and Troy D. Querec for the sample analyses in the USA and for helpful comments throughout the project.

The first author, EY, used AI-assisted tools (Microsoft Copilot and ChatGPT, OpenAI) to assist development and debugging of analysis codes in the script. However, the tools were not used to generate or interpret data, or on any scientific decisions. The code, and output of the analysis, were verified by EY.

## AUTHOR COUNTRIBUTIONS

JD: conceptualization, funding acquisition, investigation, project administration, resources, supervision, lead, validation, writing—original draft, writing—review & editing. EY: formal analyses, methodology, visualization, writing—original draft, writing—review & editing. LSAM: supervision, validation, writing—review & editing. KME: supervision, validation, writing—review & editing. All co-authors: formal analyses, methodology, writing—review & editing.

## DATA AVAILABILITY

The individual level anonymized data will be freely made available at B2Share (b2share.eudat.eu).

## ABBREVATIONS

CCCE: Center for Cervical Cancer Elimination
CIN2+: Cervical intraepithelial neoplasia grade 2 or worse, including adenocarcinoma in situ and invasive cervical cancer
Ct: Cycle thresholds
GE: Genome Equivalent
HPV: Human papillomaviruses
HSIL: High-grade Squamous Intraepithelial Lesion
IARC: International Agency for Research on Cancer
ICC: Invasive cervical cancer
IS: International Standard
IU: International Unit
LBC: Liquid-based Cytology
LSIL: Low-grade Squamous Intraepithelial Lesion
NRL: National Reference Laboratory
PAF: Population Attributable Fraction
TPP: Target Product Profile
WHO: World Health Organization

